# Low prevalence of COVID-19 Exposure is Coincident with Self-reported Compliance with Public Health Guidelines among Essential Employees at an Institute of Higher Education

**DOI:** 10.1101/2021.03.21.21251651

**Authors:** Tracy L. Nelson, Bailey Fosdick, Laurie M. Biela, Hayden Schoenberg, Sarah Mast, Emma McGinnis, Michael C. Young, Lori Lynn, Scott Fahrner, Laura Nolt, Tina Dihle, Kendra Quicke, Emily N. Gallichotte, Emily Fitzmeyer, Greg D. Ebel, Kristy Pabilonia, Nicole Ehrhart, Sue VandeWoude

## Abstract

**Importance:** Detailed analysis of infection rates paired with behavioral and employee reported risk factors are vital to understanding how COVID-19 transmission may be inflamed or mitigated in the workplace. Institutes of Higher Education are heterogeneous work units that supported continued in person employment during COVID-19, providing an excellent test site for occupational health evaluation.

**Objective:** To evaluate self-reported behaviors and SARS-CoV-2 among essential in-person employees during the first six months of the COVID-19 pandemic.

**Design:** Cross-sectional, conducted from July 13-September 2, 2020.

**Setting:** Institute of Higher Education in Fort Collins, Colorado.

**Participants:** Employees identified to be an essential in-person employee during the first six months of the pandemic (1,522 invited, 1,507 qualified, 603 (40%) completed the survey). Of those completing the survey, 84.2% (508) elected to participate in blood and nasal swab sample collection to assess active SARS-CoV-2 infection via qRT-PCR, and past infection by serology (overall completion rate of 33.7%). Eligibility included ≥ 18 years old, able to read and understand English, not currently experiencing cough, shortness of breath or difficulty breathing, fever >100.4F (38C), chills/shaking with chills, muscle pain, new or worsening headaches, sore throat or new loss of sense of taste/smell.

**Exposure:** Self-reported COVID-19 protective behaviors

**Main Outcome(s) and Measure(s):** Current SARS-CoV-2 infection detected by qRT-PCR or previous SARS-CoV-2 infection detected by IgG SARS-CoV-2 testing platform.

**Results:** There were no qRT-PCR positive tests, and only 2 (0.39%) contained seroreactive IgG antibodies. Participants were 60% female, 90% non-Hispanic white, mean age 41 years (18-70 years). Handwashing and mask wearing were reported frequently both at work (98% and 94% respectively) and outside work (91% and 95% respectively) while social distancing was reported less frequently at work (79%) then outside of work (92%) [*p < .001*]. Participants were more highly motivated to avoid exposures out of concern for spreading to others (83%) than for personal implications (63%) [*p < .001*].

**Conclusions and Relevance:** This is one of the first reports to document that complex work environments can be operated safely during the COVID-19 pandemic when employees report compliance with public health practices both at and outside work.

## Introduction

SARS-CoV-2, the cause of COVID-19, has resulted in a global outbreak with over 26 million documented infections and over 450,000 deaths in the United States alone.^1^ The pandemic has resulted in closures of non-essential businesses nationwide as well as universities, schools, churches, restaurants, and gyms to name but a few impacted workplaces. ^1^ In the early days of the pandemic, the extent of these closures varied by state and community, as did the effects of these lock downs on COVID-19 incidence. In communities that saw dramatic drops in case numbers, considerations for safe workplace re-entry were quickly adopted. Empirically derived models that minimized the risk of outbreaks in the workplace were lacking; therefore, states, communities, and businesses were forced to improvise to develop protocols that would allow a safe return to normal workforce productivity and function.

While there have been a number of reports of workplace associated COVID-19 transmission, these have largely been associated with occupations with high exposure rates, or workplace settings where public health safety guidelines are difficult to practice (i.e. skilled nursing facilities, prisons, and meat processing plants).^2-5^ However there is a paucity of information reporting COVID-19 incidence in workplace settings where explosive outbreaks have not been confirmed. One of the most complex workplaces that has supported continued employment by so-called ‘essential employees’ are Institutes of Higher Education (IHE). IHE’s must consider learning and living environments for young adults, many away from home for their first time.

These communities represent high risk areas for disease transmission and outbreaks as they include residence halls and other congregate living spaces including dining halls, locker rooms, lecture halls and laboratories. COVID-19 diagnostic or research investigations are performed in IHE laboratories, and while strict personal protective equipment (PPE) adherence is enforced in these settings, they nonetheless represent potential exposure risk areas. IHE may also include publicly accessed facilities, such as teaching hospitals or other service centers. Despite these occupational exposure risks, IHE facilities and essential services have, for the most part, not been completely shut down during stay-at-home phases of the COVID-19 pandemic. Thus, IHEs represent a specialized heterogenous community in the context of the pandemic.

Nearly all IHE developed plans for managing student return to campus during Fall, 2020, with varying levels of success. Return to work models for IHE employees have had to consider not only student safety, but the safety of essential employees who maintain the operations and physical facilities, as well as the faculty educators, and researchers^6^, which by and large has not been closely evaluated.

This study was therefore conducted to evaluate the effectiveness of a workforce re-entry model used by one IHE to assess the risk for COVID-19 among essential workers before students returned to campus in Fall 2020. This institution included laboratories investigating SARS-CoV-2, a CLIA certified diagnostic laboratory performing SARS-CoV-2 quantitative reverse transcription polymerase chain reaction (qRT-PCR), a veterinary teaching hospital, as well as other research, teaching, and service activities that continued on campus through state ‘shut down’ orders. We specifically investigated the prevalence of SARS-CoV-2 among asymptomatic individuals as well as previous exposure to the virus. We also report on the frequency of protective behaviors at and outside the workplace as well as essential workers’ concerns over both contracting COVID-19 and exposing others.

## Methods

### Study Design and Setting

This was a cross-sectional study conducted at an Institute of Higher Education in Fort Collins, Colorado.

### Participants

Employees of Colorado State University were recruited to participate if they were identified by the human resources office to be an in-person essential employee at the time of the stay-at-home (March 26, 2020-April 26, 2020) and safer-at-home orders (April 27, 2020-July 6, 2020) implemented by the State of Colorado Governor’s Office. Initial invitations for participation in the study were sent on July 13, 2020, and the survey closed on September 2, 2020.

Participants that had not enrolled were re-invited at approximately 2-week intervals up to 3 times. Participants were first qualified through an on-line survey using Research Electronic Data Capture (REDCap), a secure, web-based software program designed to support data capture for research studies.^7-8^ Qualification included being 18 years or older, being able to read and understand English and not currently having cough, shortness of breath or difficulty breathing, fever >100.4F (38C), chills/shaking with chills, muscle pain, new or worsening headaches, sore throat or new loss of sense of taste/smell. Participants who reported these symptoms were advised to contact their primary care physician and/or seek testing at one of the local county SARS-CoV-2 testing sites.

### Procedures

The study was approved by the Institutional Review Board at Colorado State University (CSU). Participants gave their informed consent prior to enrollment. Qualified participants were directed to a 90-question survey that included questions about work environment, COVID-19 protective behaviors including social distancing, handwashing and mask wearing, past symptoms, exposures and testing as well as perceptions of risk and health behaviors. After completing the survey, individuals were scheduled for a nasal mid-turbinate swab and venipuncture at the CSU Human Performance Clinical Research Laboratory (HPCRL) located on the CSU campus.

### Participant Sampling and Sample processing

Blood and nasal swabs were collected by research staff at the HPCRL. Samples were de-identified prior to sending to laboratories for analysis. Blood samples were obtained from the antecubital vein into a serum collection tube. Research staff at the HPCRL were trained by a staff physician at the CSU Health Network on mid-turbinate nasal swab collection. Blood was centrifuged for 10 minutes at 1300 g. Serum was separated into 0.5ml aliquots and frozen at - 20°C until testing. Swabs were placed in conical tubes containing 3ml viral transport media (Hanks Balanced Salt Solution, 2% FBS, 50mg/ml gentamicin, 250ug/ml amphotericin B/fungizone) and transported for laboratory analysis.

### Serology and qRT-PCR Testing

Serum was tested for IgG antibodies seroreactive to SARS-CoV-2 antigens by the National Jewish Health CLIA Diagnostic Laboratory using an Abbott Architect IgG SARS-CoV-2 testing platform. ^9-10^ RNA was extracted from nasal swabs and qRT-PCR was performed as previously described.^11-14^ Samples with qRT-PCR reactivity were sent to a CSU CLIA certified diagnostic laboratory for validation.^15^

### Statistical Analysis

Descriptive statistics and visualizations were created using the statistical program R.^16^ All hypothesis tests were performed at the .05 level. To compare reported protective behavior usage across types and locations, chi-squared tests were performed. These tests were also used to compare participant concerns over exposing others to COVID-19 versus contracting it themselves. Mann Whitney U tests were used to assess whether protective behavior frequency (“sometimes” vs “mostly”/“always”), concern about contracting COVID-19 (“not much”/“some” vs “quite a bit”/“very”) and concern about exposing others to COVID-19 (“not much”/“some” vs “quite a bit”/“very”) varied by age. The relationships between protective behavior frequencies and concerns were measured using Spearman rank correlations and significance was assessed using permutation tests. Logistic regression, along with AIC measures of model fit and likelihood ratio tests (LRT), was used to quantify the relative effects of age and workplace on social distancing reports at work (“sometimes” vs “mostly”/ “always”).

## Results

There were 1,522 essential in-person employees invited to participate in this surveillance program; 15 did not qualify due to having one of the exclusionary symptoms, 603 (40%) of those who qualified (1507) completed the survey. Eighty-four percent of those completing the survey (508) were sampled for active infection via qRT-PCR and past infection by serology, for an overall completion rate of 33.7%. Enrolled employees represented all work units invited to participate.

Descriptive statistics for the 508 participants who completed all aspects of the study are illustrated in Table 1. Participant ages ranged from 18 to 70 years with an average age of 41 years. Sixty percent of participants were female and nearly 90% were non-Hispanic white.

**Table 1.** Demographic, health and unit information reported by study participants (n=508).

|  |  |
| --- | --- |
| **Workplace** |  |
| Veterinary Teaching Hospital | 199 (39.2%) |
| Facilities Management | 88 (17.3%) |
| Housing and Dining Facilities | 42 (8.3%) |
| Residential Dining | 16 (3.1%) |
| Health Network Medical | 14 (2.8%) |
| Laboratory Animal Resources | 12 (2.4%) |
| Other (e.g. research lab, central receiving, police, univ. housing) | 137 (27.0%) |
| **Age (years)** |  |
| 18-25 | 48 (9.4%) |
| 26-35 | 159 (31.3%) |
| 36-45 | 125 (24.6%) |
| 46-55 | 85 (16.7%) |
| 56-65 | 80 (15.8%) |
| 66-75 | 11 (2.2%) |
| **Gender** |  |
| Female | 305 (60.0%) |
| Male | 200 (39.4%) |
| Other | 3 (0.6%) |
| **Hispanic/Latino** |  |
| Yes | 46 (9.1%) |
| **Race** |  |
| White | 451 (88.8%) |
| Asian | 30 (5.9%) |
| American Indian or Alaskan Native | 17 (3.3%) |
| Black or African-American | 8 (1.6%) |
| Native Hawaiian or other Pacific Islander | 4 (0.8%) |
| Other | 8 (1.6%) |
| **BMI (kg/m2)** |  |
| Underweight (<18.5) | 9 (1.8%) |
| Normal (18.5-24.9) | 228 (46.8%) |
| Overweight (25.0-29.9) | 142 (29.2%) |
| Obese (≥30.0) | 107 (22.0%) |
| **Other** |  |
| Exercise* | 388 (76.4%) |
| Asthma | 62 (12.2%) |
| Diabetes | 11 (2.2%) |
| History of Blood Clots | 16 (3.1%) |
| High Blood Pressure | 52 (10.2%) |
| Allergies | 250 (49.2%) |

\*Number reporting yes to the question: Do you get at least 150 minutes of moderate exercise (brisk walking, slow biking, dancing) or 75 minutes of vigorous exercise (running/jogging, swimming, basketball, tennis) per week?

Participants reported few chronic conditions, except for overweight or obesity where 51% fell into this category based on self-reported height and weight.^17^

Most of the participants (80%) reported spending greater than 20 hours per week on the CSU campus since February 1, 2020. Two-thirds of participants reported spending most of their time at work with employees from the same unit (eTable 1).

Participants reported practicing protective behaviors while at work and outside work. Table 2 shows that while at work, most employees reported mostly or always wearing a face mask (98%) and frequently washing their hands (94%) while fewer reported mostly or always social distancing (79%) [*χ^{2}= 332, df = 4, p < .001 for test of differences across behaviors*]. Outside work, the percentages were similar except 92% of respondents reported practicing social distancing [*χ^{2}= 37, df=4, p < .001 for test of differences across behaviors]*. The frequency of social distancing at and outside work was significantly different [*χ^{2}= 80, df = 2, p < .001*]. Overall, protective behavior usage varied little by age, except social distancing, with younger age groups reporting less social distancing at work (eFigure 1) [*Mann Whitney U=17,223, p=*.*003*]. There were few differences by gender (eFigure 2), but bigger variation in these behaviors by workplace (eFigure 3). Considering social distancing at work specifically, we found levels of social distancing varied significantly both by workplace and age group (*X^{2}= 27*.*8, df = 6, p < .001; χ^{2}= 15*.*5, df = 5, p = .009)*, with workplace explaining more variability than age (*AIC=458*.*2 vs AIC=470*.*0*). Nevertheless, even accounting for workplace, age remains significantly associated with social distancing levels at work (*LRT p = .043*).

**Table 2.** **Protective behaviors among participants while at work and outside work by their frequency.**

|  |  | **Always** | **Mostly** | **Sometimes** |
| --- | --- | --- | --- | --- |
| **At Work** | Social distance, No. (%) | 200 (39) | 203 (40) | 104 (21) |
|  | Wash hands, No. (%) | 397 (78) | 83 (16) | 27 (6) |
|  | Facemask, No. (%) | 453 (89) | 43 (9) | 11 (2) |
| **Outside Work** | Social distance, No. (%) | 340 (67) | 125 (25) | 43 (8) |
|  | Wash hands, No. (%) | 356 (70) | 109 (21) | 43 (9) |
|  | Facemask, No. (%) | 420 (83) | 61 (12) | 27 (5) |

Note: The percentages in each row sum to 100.

Employees reported being more concerned (e.g. very to quite a bit) about exposing others to COVID-19 (83%) compared to contracting COVID-19 (63%) (See Table 3) [*χ^{2}= 49, df = 1, p < .001*]. The concerns for contracting COVID-19 appear to vary by age with older adults being most concerned [*Mann Whitney U=28,624, p=*.*51*]; there was little variation by age in concern for exposing others, with most people reporting being very concerned (eFigure 4) [*Mann Whitney U=20,164, p=*.*08*].

**Table 3.** **Participant concerns for contracting and exposing others to COVID-19 by their frequency.**

|  | **Very** | **Quite a bit** | **Some** | **Not much** |
| --- | --- | --- | --- | --- |
| Concern for contracting COVID-19, No. (%) | 147 (29) | 172 (34) | 120 (24) | 66 (13) |
| Concern for exposing others to COVID-19, No. (%) | 308 (61) | 111 (22) | 69 (14) | 17 (3) |
| Important to know if previously exposed, No. (%) | 236 (47) | 127 (25) | 127 (25) | 16 (3) |

Note: The percentages in each row sum to 100.

Not surprisingly, we found that protective behaviors varied by concern for contracting COVID-19 Those who reported always social distancing, washing their hands and wearing a facemask both at work and outside work were more concerned with contracting COVID-19 than those who reported these behaviors some of the time (Figure 1; Spearman rank correlations ranged 0.10 - 0.24). Similarly, we observed protective behaviors were practiced more commonly by individuals with significant concerns about exposing others to COVID-19 (Figure 2; Spearman rank correlations ranged 0.05 - 0.20), though social distancing at work was not significantly correlated with concern for exposing others.

**Figure 1.**
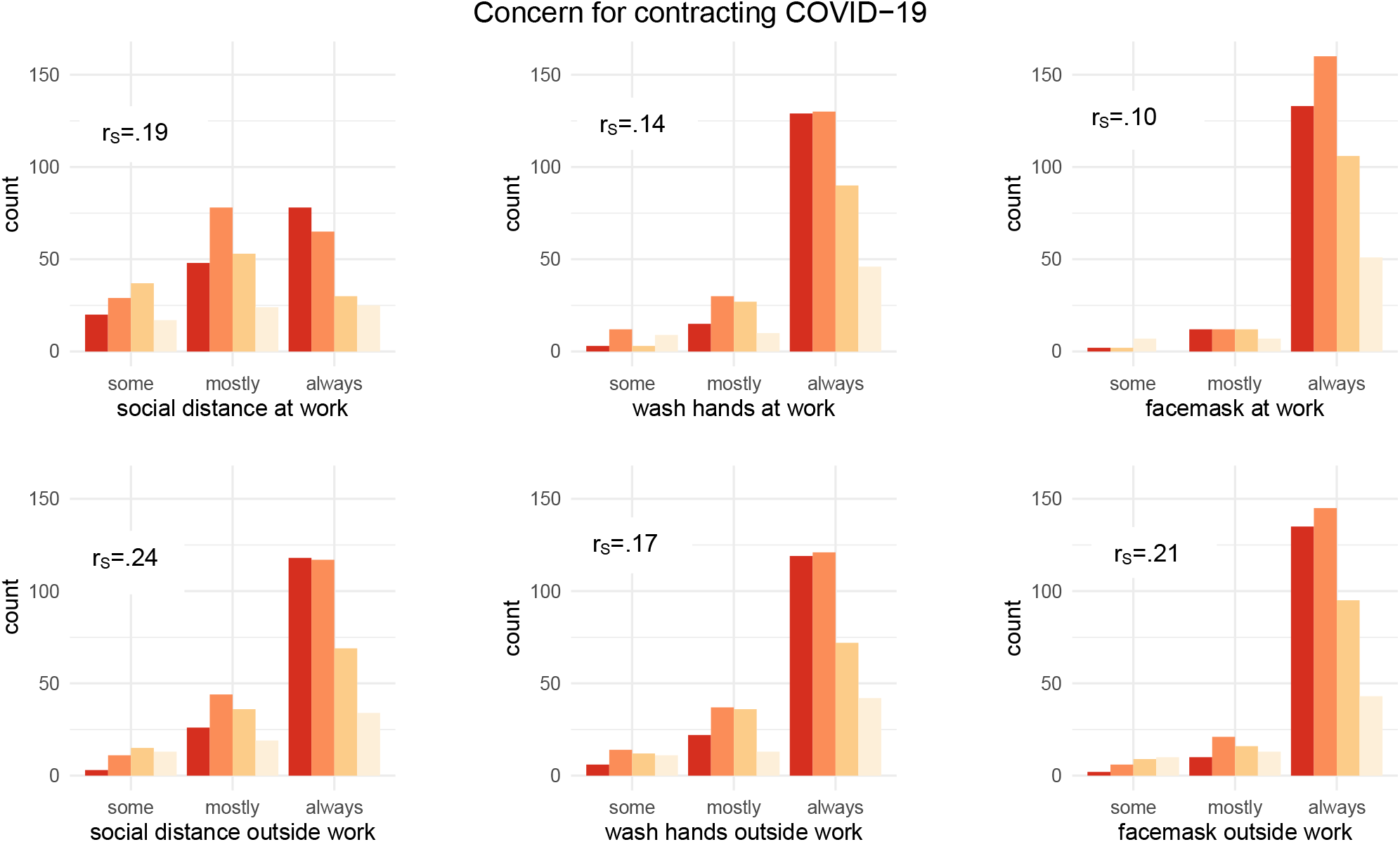
Frequency of protective behaviors by level of concern for contracting COVID-19. Dark red bars indicate high level of concern and correlate with reported compliance with protective behaviors. All Spearman rank correlations (r_s_) significant at 0.05 level.

**Figure 2.**
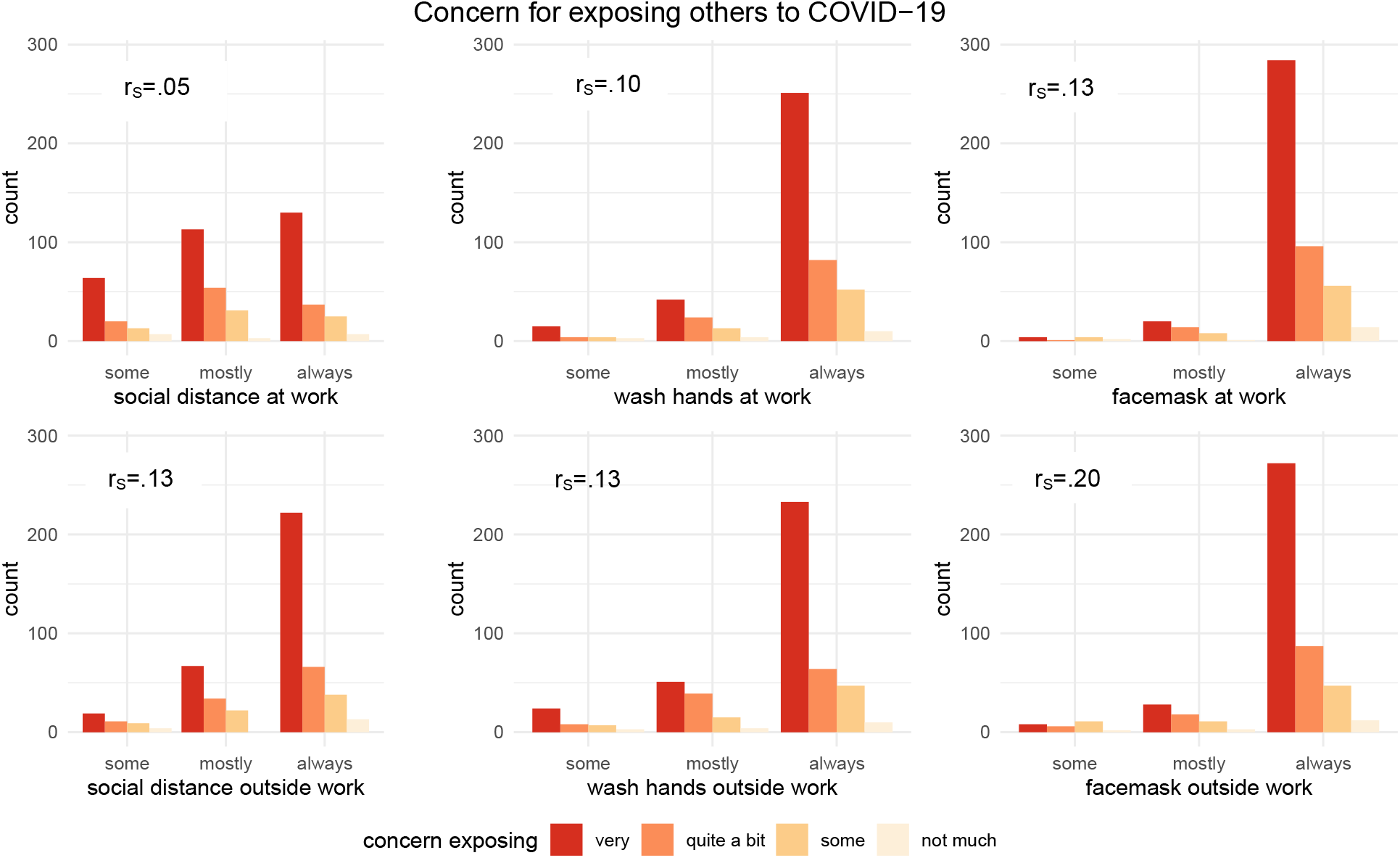
Frequency of protective behaviors by level of concern for exposing others to COVID-19. Dark red bars indicate high level of concern and are correlated with protective behaviors. All Spearman rank correlations (r_s_) significant at 0.05 level, except social distancing at work.

### Nasal Swab and Serologic test results

Five hundred and eight nasal swabs were tested using qRT-PCR. Five hundred and two results were clearly negative. Six results were inconclusive in a laboratory-based screening test and were sent to the CSU CLIA certified diagnostic laboratory for confirmatory testing where all tests were confirmed negative (0 positive qRT-PCR for 508 tests). Two serum samples out of 508 tested seroreactive for IgG antibodies, indicating previous SARS-CoV-2 exposures (0.39%). Of the two seropositive workers, one reported having a previous positive qRT-PCR test on the de-identified survey, the other individual had not been previously tested but reported previous COVID symptoms.

Overall, most employees reported no known contact with positive cases (92%) or contact with a symptomatic individual (87%). Nearly 13% reported having had a previous qRT-PCR test, and 4.5% reported a previous serology test. Despite the low rates of previous testing or positivity, 28% of participants reported having had previous COVID-like symptoms (eTable 2).

## Discussion

The Occupational Health and Safety Administration (OSHA) has stratified workplace exposure into four risk categories.^2^ Occupations and workplace environments at high or very high risk include those with exposure to known or suspected sources of virus, including health care and morgue workers, and laboratories handling live virus. Jobs with frequent close contact to people in high-density work environments, such as schools and retail environments have been classified as medium risk. Specific occupations in the US that have reported high levels of exposure have included skilled nursing facilities, workers in meat and poultry processing plants, and prison staff.^2-5^ The vast majority of workplace situations have not been closely evaluated for exposure to SARS-CoV-2. IHE represent a heterogeneous setting to test exposure of a diverse population of employees who reported to work during the first six months of the COVID-19 pandemic.

The first step in our IHE workforce re-entry model included inviting all non-symptomatic essential workers for SARS-CoV-2 testing using qRT-PCR. Our goal was to identify asymptomatic workers and temporarily remove them from the workforce. We did not identify any positive cases among this asymptomatic group of employees. Further, we found that only two participants were previously exposed despite 143 reporting COVID-19 symptoms in the past. This very low rate of seroprevalence (0.39%) was nearly 10% of the approximate rate of seroprevalence (3.6%) within the county at the time of this study. ^18^, indicating a far lower rate of exposure among CSU employees than other persons living in Larimer County.

The absence of any positive cases and very low levels of seroprevalence despite most of these employees working on campus > 20 hours per week, as well as 8% reporting prior contact with someone who tested positive, are quite remarkable (eTable 2). These numbers are likely attributed to the high percentage of employees who reported participating in protective behaviors, and generally reported responsibility for protecting their own health and that of others (Figures 1 and 2). We found that most employees (>90%) reported regularly washing their hands and wearing a face mask (Table 2). Social distancing was also reported by a high percentage outside of work (92%) (Table 2). Our findings correlate with reports that student behaviors were most responsible for spread of SARS-CoV-2 on campuses, and with proper precautions, classrooms and other formal spaces were not hot spots for virus spread. ^19^ Importantly, these findings indicate that IHE can safely operate when employees responsibly practice public health guidelines for infection control both at and outside of work.

As noted previously, a smaller but still substantial percentage of employees reported social distancing at work (79%) (Table 2). This suggests that work conditions may not always be conducive to practicing social distance behaviors, and we did find differences in social distancing practice by work unit (eFigure 3).

We also found social distancing to vary by employees age, with 100% of those > 65 years old reporting high levels of social distancing at work compared to about 70% of those 18-25 years old (eFigure 1). It is not surprising that those with higher vulnerability to severe disease were more likely to conform to social distance recommendations. Since mask wearing and hand washing are independently controlled and more easily managed during workplace interactions, these behaviors would tend to be easier compliance measures than social distancing. Workplace explained more of the variation in social distancing than age, however, suggesting there may be workplace situations where social distancing is not always possible.

Adherence to protective behaviors both in the workplace and at outside of work correlated with reported concerns for contracting COVID-19 (Figure 1). Individuals who reported always social distancing, washing their hands and wearing a facemask at work and outside work also reported being most concerned with contracting COVID-19. These findings are similar to those found among a U.S. sample during the first week of the pandemic, where social distancing and handwashing were most predicted by perceived likelihood of becoming infected. ^20^

Interestingly, employees were more concerned about exposing others to COVID-19 than contracting COVID-19 (83% vs. 63%) and this concern translated to protective behaviors (Figure 2). Such concern can be described as prosocial, or behavior that is helpful and intended to promote social acceptance. ^21^ A recent study conducted in Sweden found those scoring higher on a prosociality measure were more likely to follow physical distancing guidelines, stay at home when sick and buy face masks during the COVID-19 pandemic. ^22^ There are multiple drivers of prosocial behavior including empathy, as well as building a sense of shared social identity which can be done through effective leadership. ^23^ Prosocial behavior has shown to lead to greater positive affect, meaningfulness, empathy and social connectedness ^24^, which may then not only influence employees desire to protect their co-workers from an infectious virus (or any other threat), but also overall morale.

Limitations of this study include the use of self-reported data which could have resulted in response bias for reported protective behaviors. This was a cross-sectional study so we could not determine temporality between protective behaviors and COVID-19 outcomes; however, given that there were not any positive cases and only two participants who were seropositive for COVID-19 suggests previous exposure unlikely influenced protective behaviors, and that protective behaviors contributed to the low level of disease in this population. There may have also been nonresponse bias, whereby those who were invited and did not participate differed in some way from those who did. However, our participation rate (33.7%) and number of respondents (n=508) was significant and helps to mitigate this concern.

## Conclusion

This study reported extremely low levels of SARS-CoV-2 active or previous exposures in a large cohort of employees whose job type required them to work on campus during COVID-19 state restrictions. Our findings suggest that the absence of positive cases and low seroprevalence (only 10.8% of the seroprevalence rate in Larimer County at this time) are likely associated with the high rates of protective behaviors by employees at work, and very importantly, outside of work. Although these results reflect the experience at one Institute of Higher Education, we believe these results could be generalized to other Institutes of Higher Education as well as other complex work environments.

Future studies need to consider the important factor of practice of protective behaviors outside the workplace, as no matter how tightly social distancing, mask wearing and washing hands are regulated or expected at work, COVID-19 exposure occurring outside of work can enhance risk of spread in the workplace. Further, prosocial behavior appears to be a strong motivator to practice safe behaviors and should be fostered by unit and institutional leaders as it may help such institutions successfully remain open.

## Data Availability

Data can be made available upon request.

## Acknowledgments

We would like to acknowledge Mary Nehring for her assistance with sample transport as well as staff at the Larimer County Department of Health and Environment for their assistance with the survey.

