## Supplemental Tables 1-2 and Figures 1-4 for "Low prevalence of COVID-19 Exposure is Coincident with Self-reported Compliance with Public Health Guidelines among Essential Employees at an Institute of Higher Education"

### eTable 1: Participants reported contact with others while at the workplace, ranging from none of the time to ≥50% of the time.

| Contact in the workplace | None | <50% | ≥50% |
|---|---|---|---|
| Employees, same unit, No. (%) | 10 (2) | 163 (32) | 335 (66) |
| Employees, outside unit, No. (%) | 141 (28) | 320 (63) | 47 (9) |
| General public, No. (%) | 193 (38) | 286 (56) | 29 (6) |
| Students, No. (%) | 169 (33) | 263 (52) | 76 (15) |

Note: The percentages in each row sum to 100.

### eTable 2: Participants reported history of exposure, testing and symptoms for COVID-19.

| | Yes | No | Don't Know |
|---|---|---|---|
| Contact with COVID-19 positive individual, No. (%) | 41 (8.1) | 299 (58.9) | 168 (33) |
| Contact with individual showing symptoms, No. (%) | 68 (13.4) | 295 (58.1) | 145 (29) |
| Previous PCR test for COVID-19, No. (%) | 65 (12.8) | 443 (87.2) | |
| Previous Positive PCR test, No. | 1 | | |
| Previous Serology test for COVID-19 antibodies, No. (%) | 23 (4.5) | 485 (95.5) | |
| Previous Positive Serology test, No. | 1 | | |
| Previous COVID-like symptoms, No. (%) | 143 (28.1) | 365 (71.9) | |

**eFigure 1: Frequency of protective behaviors at and outside the workplace by age.**

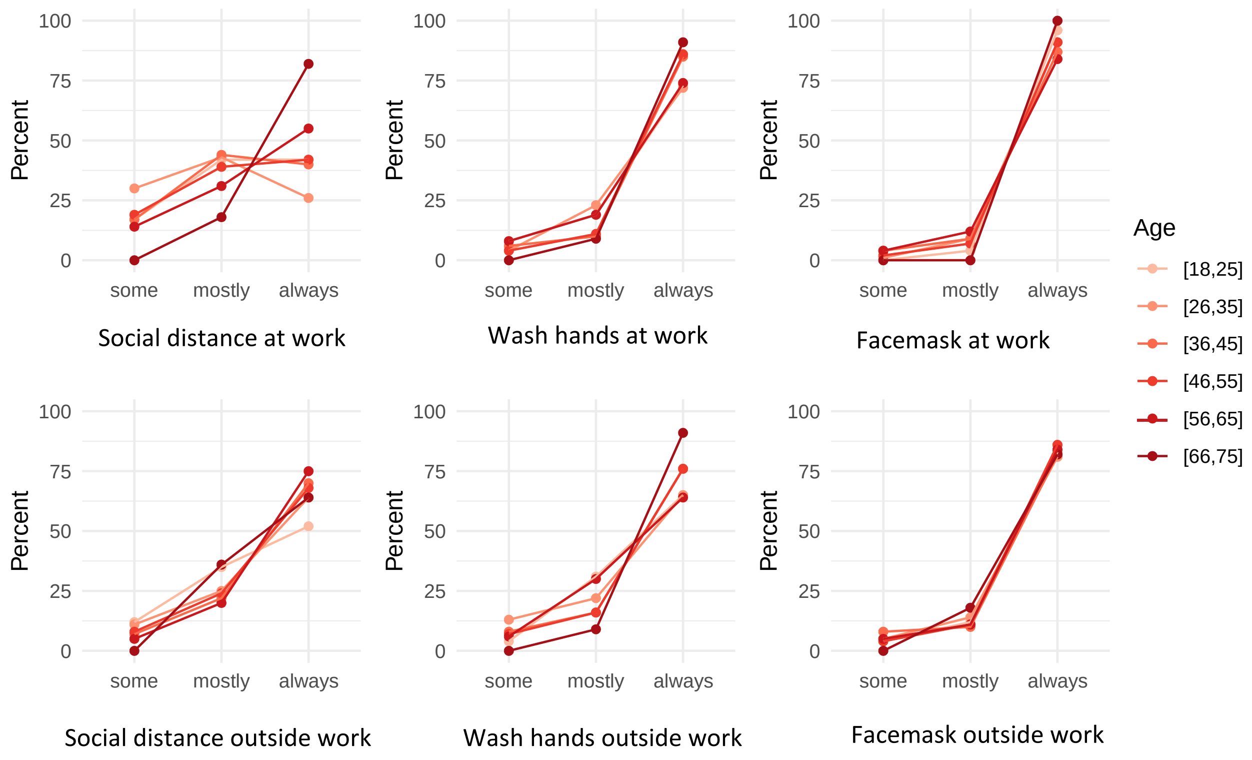

**eFigure 2: Frequency of protective behaviors at and outside the workplace by gender.**

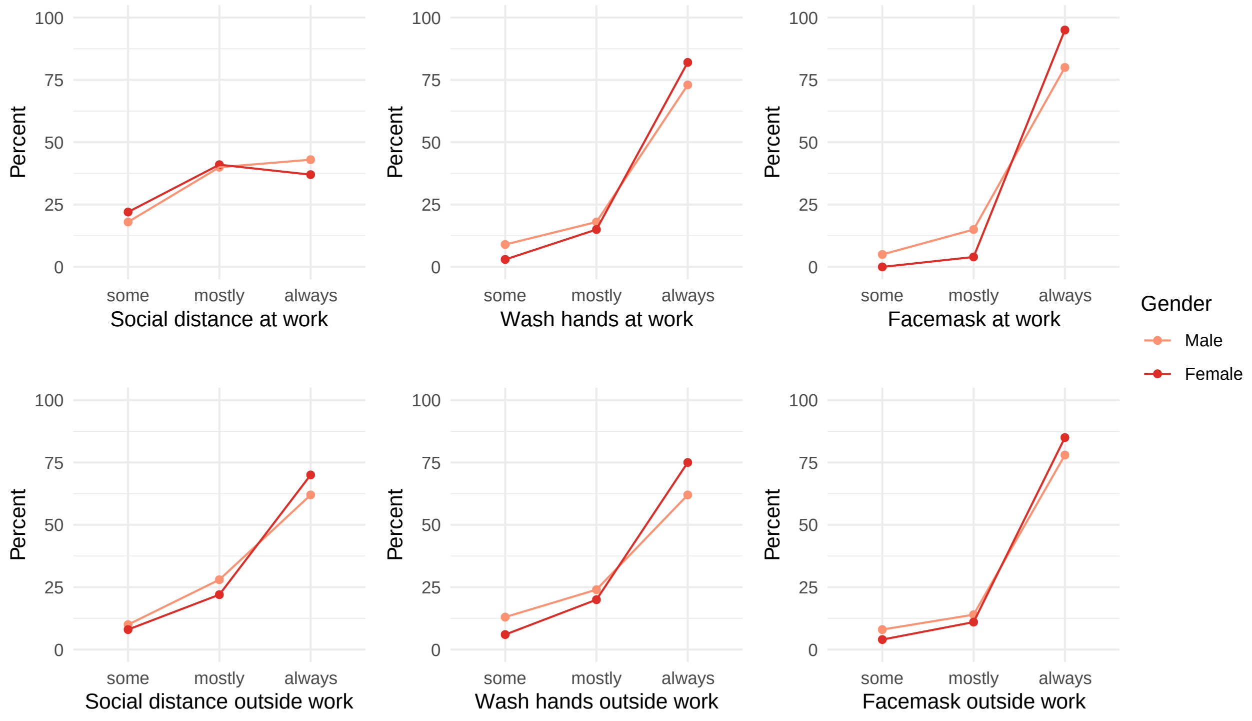

**eFigure 3: Frequency of protective behaviors at and outside the workplace by work unit.**

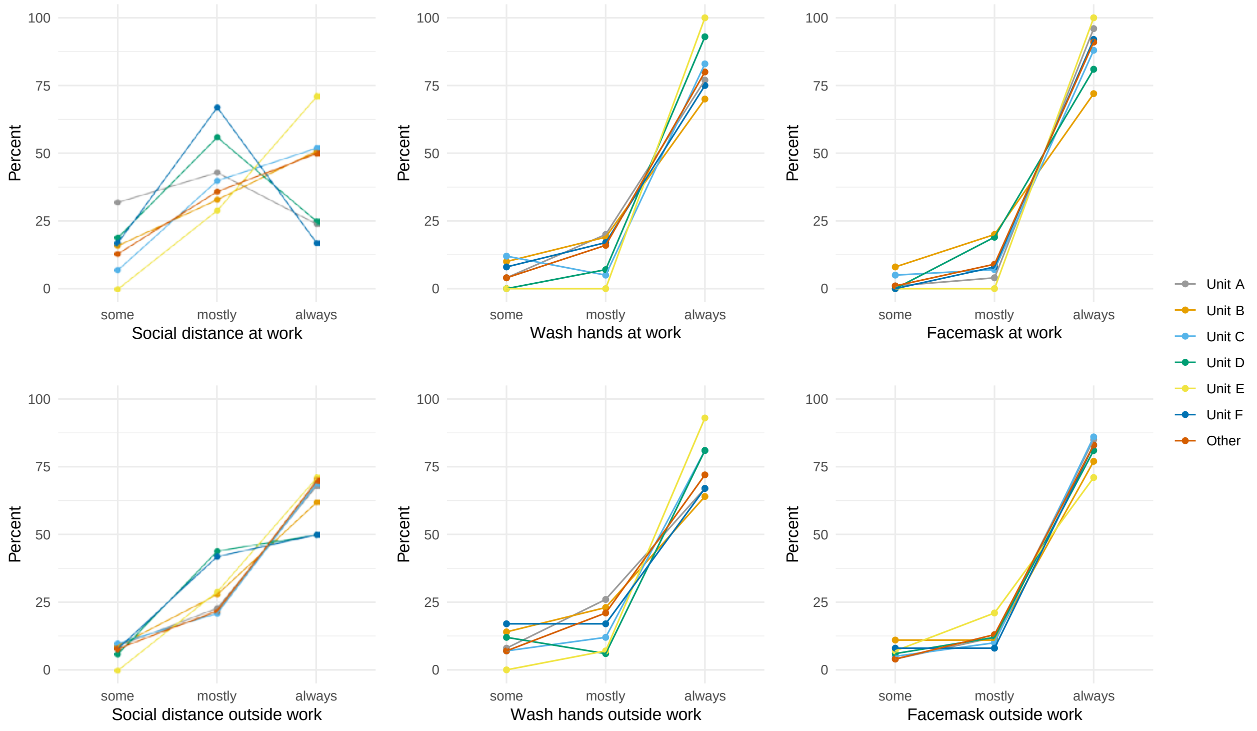

**eFigure 4: Concerns for contracting and exposing others to COVID-19 by age.**

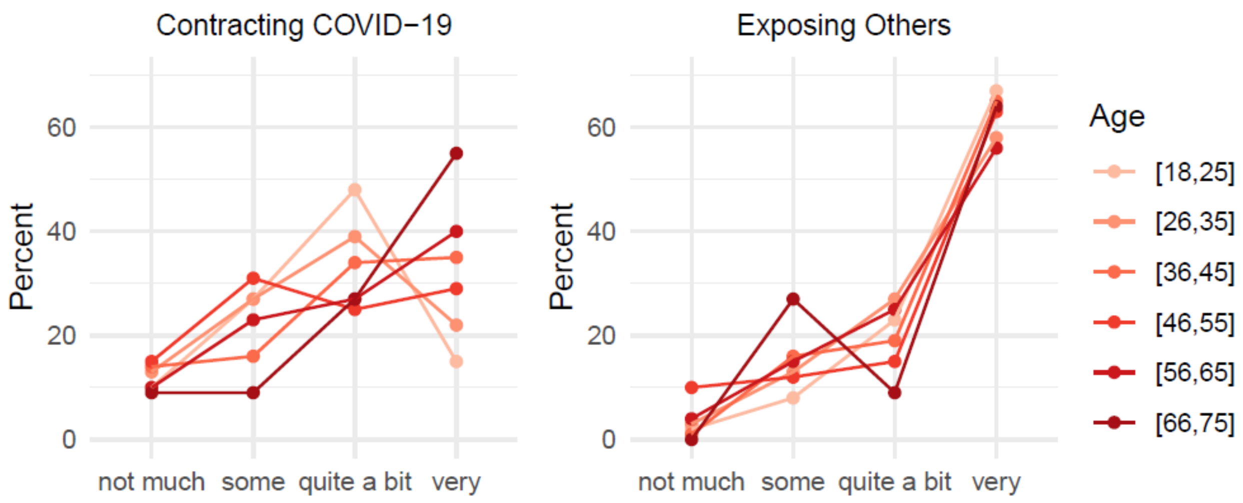
